# Air recirculation role in the infection with COVID-19, lessons learned from Diamond Princess cruise ship

**DOI:** 10.1101/2020.07.08.20148775

**Authors:** Orouba Almilaji, Peter W Thomas

## Abstract

**Objectives:** The Diamond Princess cruise ship is a unique case because it is the place at which testing capacity has reached its highest rate in the world during the COVID-19 pandemic. By analysing data that are collected about the current COVID-19 outbreak onboard, and by considering the design of the air conditioning system of the ship and virus transmission modes on cruise ships, this study aims to raise the hypothesis regarding the role of poor ventilation systems in the spread of COVID-19.

**Design:** This is an analysis of count data that has been collected by the onboard clinic up to the 20^th^ February 2020. Symptomatic infection rates during the quarantine period in cabins with previous confirmed cases are compared to these in cabins without previous confirmed cases.

**Results:** Symptomatic infection rate during the quarantine period in cabins with previously confirmed cases is not significantly higher than that in cabins without previously confirmed cases. Age doesn’t appear to be a cofounder.

**Conclusions:** Airborne transmission of COVID-19 through the ventilation system onboard could explain the virus spread into cabins during the quarantine period.

**Strength and Limitations of the study:**

- The study described an important intervention measure that is ignored by national authorities to reduce exposure and limit the transmission of COVID-19 in confined places.
- The study reviewed the Diamond Princess cruise ship deck plans, the design of its HVAC (heating ventilation air-conditioning) system, and previous viral outbreaks on cruise ships.
- The study considered all other studies that highlighted the airborne transmission mode of COVID-19.
- The study only considered the airborne transmission mode to explain the infection with COVID-19 in passengers’ cabins on the Diamond Princess cruise ship.
- The study used published count data to calculate the infection rates.

## Introduction

Unreported cases, and limited testing capabilities for the SARS-CoV-2 have led to uncertainty about the actual number of people that are infected, died, or recovered from COVID-19 around the world. As most of the people on Diamond Princess (DP) cruise ship have been tested for COVID-19, the DP ship has become an important case study for research about the COVID-19 outbreak.

Because of 10 lab-confirmed cases (CCs) of COVID-19 on the 4^th^ of February, DP has been quarantined for the period of 5th-19^th^ of Feb 2020. A total of 3711 individuals were on board at that first day of quarantine (2666 passengers and 1045 crew members) [1]. Crew members were provided with personal protective equipment (PPE) and instructed to follow international guidance on Infection prevention and control. Passengers were given thermometers and instructions for self-monitoring of body temperature and requested to stay in their cabins and to call if they had a fever above 37.5°c [1]. Passengers who developed fever were tested for COVID-19. If their tests confirmed positive, they disembarked the ship and isolated. Then their cabinmates -if any-consequently tested. If they are found to be positive cases too, they disembarked the ship and isolated. Otherwise, they remained on board.

By the 20^th^ of Feb, 619 COVID-19 positive cases were confirmed. Out of these cases, a total of 301 (49%) cases were symptomatic. Among these cases, there were 163 cases with recorded symptom onset dates during the quarantine period (QP) [1]. Among these 163 cases, 115 were passengers, and 48 were crew. On the 20^th^ February, over 1600 non-case, mainly passengers had disembarked the DP ship. Criteria for disembarkation of non-cases included; completion of a 14-day period without sharing a cabin with a CC, a negative test result, and no symptoms [1].

Despite the fact that substantial inactive infection of SARS-CoV-2 could have occurred prior to the QP, or during the QP due to close contact with a CC(s) within the same cabin, many new cases with recorded symptom onset dates during the QP have occurred in cabins with no previous CCs of COVID-19 and in single-occupancy cabins.

It is not possible to infer exactly when all cases on DP were infected due to the variance in COVID-19 incubation periods among individuals and to the lack of symptom presentation in many cases. However, we assume that; firstly, the infection risk is higher when there is a close contact with a sick person within a small space such as ships’ cabins. And secondly, the mean incubation period for COVID-19 is around 7 days (mean incubation period for COVID-19 is estimated to be 6.4 days [2], median incubation period is estimated to be 5.1 days [3]).

This study aims to test the null hypothesis that the infection rate of new CCs with recorded symptom onset dates during the QP in cabins with previously CCs of COVID-19, is higher to than that in cabins with no previously CCs. To eliminate the effect of pre-QP exposure to the virus as much as possible, infection rates of new CCs with recorded symptom onset on or after the median quarantine day (day 7) will be compared also. We will investigate whether dissimilar age distributions among the cabins could had been the reason for any difference.

## Methods

### Data

This is an analysis of count data published on 21^st^of Feb 2020 and collected on the DP by the ship’s onboard clinic [1]. The data in this paper comes from two tables published within a field briefing that was prepared by the Japanese national institute of infectious diseases [1]. The first table includes cases per date of onset (quarantine day), number of additional CCs of passengers in cabins with previous CCs, number of additional CCs of passengers in cabins without previous CCs, and number of additional CCs of crew members, for a total of 163 symptomatic CCs with recorded symptom onset during the QP. Data were published only for the period of 6-17^th^ of Feb. The second table was recorded on 20^th^ February and organized according to 10 age group, each with a span of 10 years. For each age group, data included population aboard, total number of CCs, and number of symptomatic CCs. In this study, symptomatic infection rate with recoded symptom onset (SIRR) equals the number of new CC with recorded symptom onset date (during the QP, or on and after the QP median day) divided by the population at risk up to the QP median day, or up to 20^th^ of Feb.

Statistical Analysis: analysis started by merging any age group that was found to be less than 5 people with the closest age group to it. Chi-square test was used to check whether symptomatic, asymptomatic, non-case counts differ by ages. If a significant difference is found, relative contribution of each cell to the total chi-square score were used to check where the differences actually lie. Two-proportions z-test (one-tailed) was used to compare SIRR in cabins. The significance level for all tests is 0.05. R (version 3.6.1) was used to run the statistical analysis. As this study was secondary retrospective analysis of anonymised published count data, formal Research Ethics approval was not required. Also, no patient and involved in this study.

## Results

By using only the passenger count data from the first prepared dataset of 115 symptomatic CCs with symptom onset dates during the QP of 6th to 17^th^ of Feb (Table 1), analysis shows that 20% occurred in cabins with a previous CC, and 80% occurred in cabins without a previous CC.

**Table 1.** COVID-19 symptomatic confirmed cases with known onset dates 6 – 17^th^ Feb 2020

| Onset dates | New CC |  |
| --- | --- | --- |
|  | In cabins with previously CC in the same cabin | In cabins without a previously CC in the same cabin |
| 06/02/2020 | 1 | 14 |
| 07/02/2020 | 4 | 25 |
| 08/02/2020 | 1 | 16 |
| 09/02/2020 | 6 | 13 |
| 10/02/2020 | 3 | 4 |
| 11/02/2020 | 3 | 8 |
| 12/02/2020 | 0 | 5 |
| 13/02/2020 | 4 | 5 |
| 14/02/2020 | 1 | 1 |
| 15/02/2020 | 0 | 1 |
| 16/02/2020 | 0 | 0 |
| 17/02/2020 | 0 | 0 |

Infection rate of new CC with recorded symptom onset dates (SIRR) were calculated as follows:
in cabins without previous CC in the same cabin:
Number of non-case passengers who did not share a cabin with a CC was 1600 on 20<sup>th</sup> Feb
Number of new CCs in cabins without previously CC during the whole QP: 92
Population at the start of the QP: 1600 + 92=1692, thus
SIRR for the whole QP is 92/1692 (**5.4%**)
Number of new CCs in cabins without previously CC during the period of on and after the median QP day: 12
Population at risk at the QP median day: 1600 + 12 = 1612
SIRR for the period of on and after the median QP day is 12/1612 (**0.7%**)

Difference in SIRRs for the whole QP was -3.1% (95% Upper CI: -1.9%), p<0.0001. Difference in SIRRs on and after the median QP day was; -0.3% (95% Upper CI: 0.2%), p=0.2.

Using the second prepared dataset (Table 2), and by excluding children and young people under 20 years of age due to small sample sizes, combining the 80-89 and 90-99 age groups, and dividing the sum of CC by the population at risk per age group, analysis showed that the highest infection rate was in the age group of “80-99” (23 %) (Figure 1).

**Table 2.** distribution of cases per age group

| Age group | Symptomatic CC | Asymptomatic CC | None-cases |
| --- | --- | --- | --- |
| 20-29 | 25 | 3 | 319 |
| 30-39 | 27 | 7 | 394 |
| 40-49 | 19 | 8 | 307 |
| 50-59 | 28 | 31 | 339 |
| 60-69 | 76 | 101 | 746 |
| 70-79 | 95 | 139 | 781 |
| 80+ | 29 | 25 | 173 |

**Figure 1.**
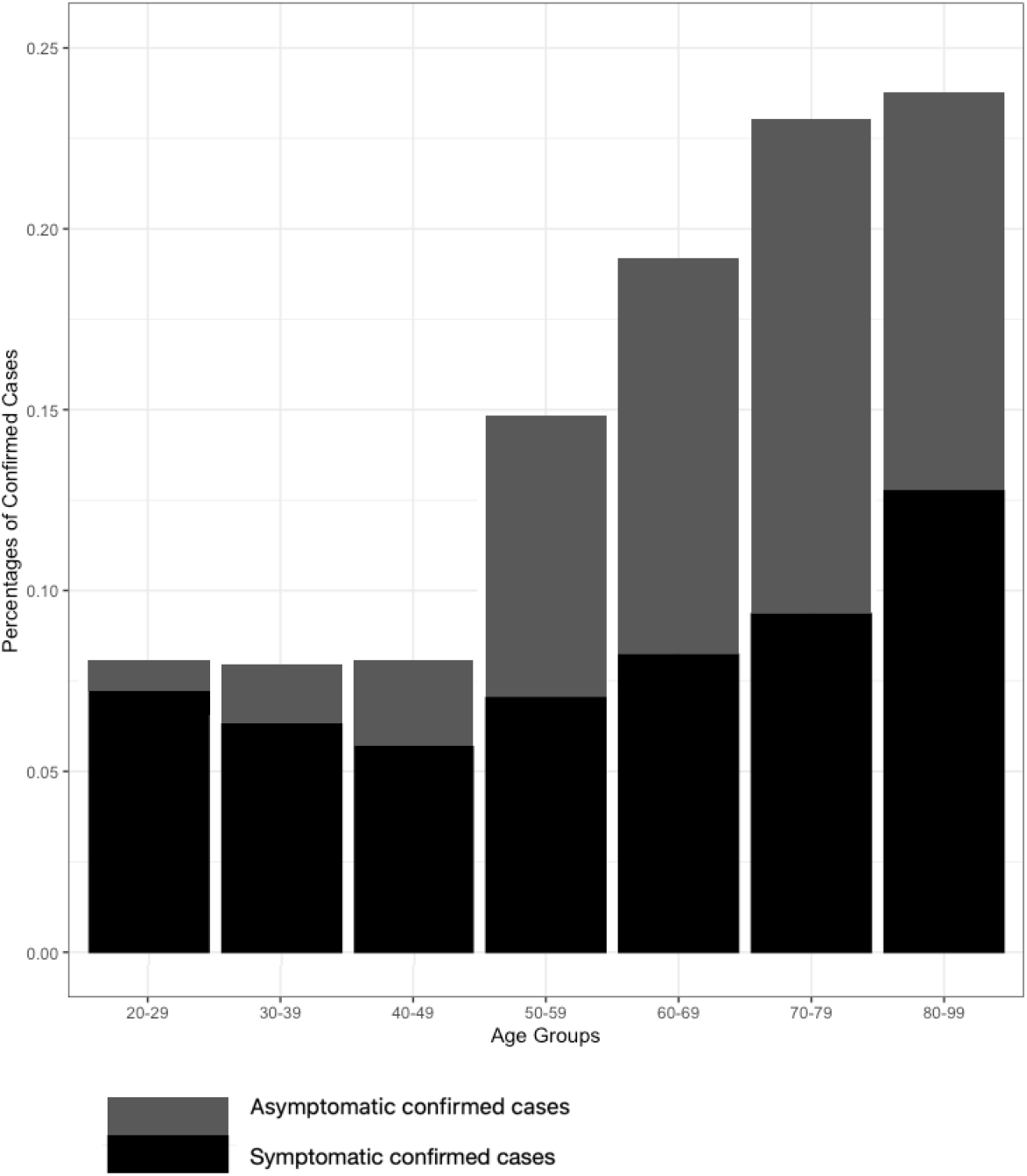
infection rates per age group

Chi-squared test showed that cases distribution differs significantly by age group (df=12, P <0.0001). The cells that contribute most to the chi-square statistic are asymptomatic CC in the age groups: “70-79”, “20-29”, “30-39”, “40-49 in descending order of contribution. These cells contribute to about 71 % to the total chi-square score and thus account for most of the difference. The symptomatic CCs contribute only to about 10% of the difference.

## Discussion

Results showed that SIRR in cabins with previously CC was not significantly higher than that in cabins without previously CCs during the whole QP. No evidence was found to conclude that SIRR in the cabins with previously CC was not significantly higher than that in cabins without previously CC on and after the median QP day. Results showed also that the counts of symptomatic cases were not dependent on age. Accordingly, a hypothetical difference in age distributions among the cabins cannot explain these results.

Due to its urgency, most of recent research has focused on the COVID-19 outbreak on DP shipboard and overlooked previous infection outbreaks on the same or other ships. Understanding the transmission modes of previous outbreaks on cruise ships could help to understand how COVID-19 infection has occurred in cabins with no reported previous cases or in a single-occupancy cabins during the QP.

In fact, outbreaks of viral infections are commonly reported on cruise ships. On DP, there was another outbreak of norovirus in 2016 [4]. Ruby Princess which is another cruise ship that has 576 cases of COVID-19 in 2020 [5], also had previous norovirus outbreaks in 2012 [6]. Other cruise ships which have witnessed COVID-19 in 2020 such as Westerdam, and Voyager of the Sea [6] had also witnessed norovirus outbreaks in the past. Though norovirus is the most common outbreak disease on cruise ships, it is not the only illness that is common on cruise ships. Legionella, Enterotoxigenic E. coli (ETEC), Campylobacter, C. perfringens enterotoxin, Shigella, Salmonella, Enterobacter, and Entameoba histolytica have also common outbreaks on cruise ships [6].

Researchers have always warned about the potential for cruise ship viral outbreaks [7, 8, 9, 10, 11]. Large number of people sharing facilities, same stocks of food and water, and air conditioning systems [10]. Such close and crowded quarters promote transmission of airborne pathogens [10, 12]. One study has found that how soon passengers boarded the ship presenting the largest determinant of their respiratory illness risk if the ship was infected [8].

Viruses onboard can be easily transmitted from one person to another by inhalation of air that contains aerosols or droplets from the infected person who cough or sneeze [13]. Airborne transmission mode plays a significant role on cruise ships [7]. Airborne transmission happens when viruses are carried by dust or droplet nuclei suspended in air [14]. Airborne dust can be resuspended by air currents after settling on any surface and droplet nuclei could remain suspended in the air for a long time and may travel long distances [14]. A new study found that COVID-19 was detectable in aerosols for up to three hours [15].

Actually, any enclosed space with dense population such as cruise ships, aircrafts, hospitals, etc, is susceptible to airborne transmission [10, 11, 16, 17, 18]. One study has found evidence to support the airborne transmission of Middle East respiratory syndrome coronavirus (MERS-CoV) in hospitals [20]. In another confined setting, airborne transmission of severe acute respiratory syndrome (SARS) has been shown in aircraft [19 -20].

Besides infectious persons, airborne transmission can be released from heating, ventilation, and air conditioning (HVAC) systems [22]. A systematic review has demonstrated strong evidence about the association between ventilation, airflow in buildings and the transmission of infectious diseases such as tuberculosis, influenza, and SARS [23]. The study showed that air flow and ventilation can affect how diseases spread indoors with the more stagnant the air is, the more likely diseases are to spread [23].

In an outbreak of COVID-19 in a restaurant in Guangzhou, China, transmission was prompted by air-conditioned ventilation and the key factor for infection was the direction of the airflow [18]. One of the recommendations from this study [18] to prevent the spread of the viruses was to improve ventilation. Another study has confirmed that there is a substantial probability that normal speaking can cause airborne COVID-19 transmission, in which speech droplets can remain suspended for tens of minutes or longer and are eminently capable of transmitting disease in confined spaces [24]. These speech droplets were shown to be influenced by the air flow and dominated by the ventilation rate [24].

A study that analysed two COVID-19 outbreaks on buses and places of worship has strongly suggested that large community gatherings, especially those in enclosed settings with minimal air ventilation, should be limited [25]. The same study [25], has suggested that the conditioning system on a re-circulating mode may have facilitated the spread of COVID-19 virus in these outbreaks. Similar to our results, the study [25] showed that passengers sitting closer to the index case on the exposed bus did not have statistically higher risks of COVID-19 as those sitting further away. The study also showed that all passengers sitting close to a window remained healthy which may be due to airflow.

Another study [26], has emphasised the importance of national authorities acknowledging the reality of airborne transmission mode of COVID-19, and recommend that adequate control measures should be implemented to prevent further spread of the COVID-19. The study [26] quoted another study [27] when mentioning all the possible precautions that should be taken against airborne transmission in indoor setting. These precautions include increased ventilation rate, using natural ventilation, avoiding air recirculation, avoiding staying in another person’s direct air flow, and minimizing the number of people sharing the same environment [27].

On airplanes, proper ventilation and high-efficiency particulate (HEPA) filters have shown to help reducing airborne transmission [11, 28]. In cruise ships, researchers have concluded that the higher the ventilation rate, the lower the number of new ill cases would be [7]. In a simulation study, HEPA filters and ultraviolet germicidal irradiation (UVGI) devices in ventilation systems were the most effective measures to control influenza on cruise ships [7]. They are shown to be more effective than masks worn by crew members and quarantining procedures [7].

The HVAC (heating ventilation air-conditioning) filtration system on the DP ship is confirmed to be comparable to those used by land-based hotels, resorts and casinos [29]. HVAC in general commercial and industrial spaces such as hotels usually have a minimum efficiency reporting value (MERV) of 3000 to 10000 nm, in which MERV refer to the effectiveness of air filters in HVAC [30]. Since SARS-CoV-2 was found to have a diameter of approximately 60– 140 nm [31], this means the ventilation system on the DP cruise ships was completely unable to filter viruses as small as SARS-CoV-2 virus. The design of the air conditioning system in DP [32, Figure 2] shows that air from different passengers’ cabins gathered in one air duct, mixed with some fresh air (30% only) and then re-supplied to the different cabins again. Moreover, DP’ deck plans showed that about half of the cabins onboard have no access to fresh air as they are either interior cabins or cabins with windows that are designed not to be opened [33]. Given that COVID-19 outbreak on DP has happened in the winter with an estimated outside temperature of -5°C [32], this means passengers on DP were fully dependent on the HVAC onboard. Since it is ordinary practice for cruise ships to mix outside air with inside air to save energy [32], then DP air conditioning system could had carried SARS-CoV-2 virus from one cabin to another. Such a probable airborne transmission mode of SARS-CoV-2 virus on the DP ship could, at least partially, explain why the infection rate in cabins with previous CC was not significantly higher than that in cabins without previous CC (and also the infections with COVID-19 in single-occupancy cabins) during the QP.

## Conclusion

There is accumulating evidence of COVID-19 spreading widely in confined settings such as restaurants, hospitals, care homes, shops, gyms, public transport, offices, schools, prisons, etc. Ventilation system design, filters, and upgrades; natural ventilation (just using outside air and not recirculating it); and airflow (direction/speed) should be all considered and evaluated when deciding what intervention measure(s) is appropriate to reduce exposure and limit the transmission of COVID-19 in a confined setting. Keeping 2 meters distance between customers in a shop, for instance, is not an effective measure without considering the air flow inside the shop. Asking older people to self-isolate inside their rooms in a care home is not an effective measure if the ventilation system in the care home does not have highly-efficient filters. Advising workers to observe distancing is not enough if the air inside the workspace is kept recirculated.

It is important to note that this is an exploratory study and it has different limitations. First and foremost, the incompleteness and quality of the published data that is used in this study in which only summary data were available, data on different cofounders were not available (such as the sex distribution among cabins), and the fact that some counts were approximate when reported such as the “1600” number which is reported as “over 1600” in [1]. Finally, though we are not discounting other important transmission modes such as close-contact droplets and fomites, in this study, we only considered the airborne transmission mode to explain the infection with COVID-19 in passengers’ cabins during the QP.

## Data Availability

No additional data available

https://www.niid.go.jp/niid/en/2019-ncov-e/9417-covid-dp-fe-02.html

## Statements

### Contributorship

OAM conceived this study. OAM, and PT designed the study. OAM analyzed the data and drafted the initial manuscript. All authors made significant contributions to the subsequent revision of the paper.

## Acknowledgments

We would like to acknowledge Doctor Jonathon Snook; a consultant at Poole hospital for his significant contribution in the study design. And we would like to acknowledge Professor Brendan Murphy, in the Mathematics and Statistics school, University College Dublin (UCD), for his kind review of the manuscript.

## Data Sharing Statement

No additional data available.

